# SARS-CoV-2 infections in children and adolescents with rheumatic musculoskeletal diseases – data from the National Pediatric Rheumatology Database in Germany

**DOI:** 10.1101/2021.03.28.21254496

**Authors:** Claudia Sengler, Sascha Eulert, Martina Niewerth, Kirsten Minden, Gerd Horneff, Jasmin B. Kuemmerle-Deschner, Caroline Siemer, Rainer Berendes, Hermann Girschick, Regina Hühn, Michael Borte, Anton Hospach, Wolfgang Emminger, Jakob Armann, Ariane Klein, Tilmann Kallinich

## Abstract

**Objectives:** Due to their underlying disease as well as therapeutic immunosuppression, children and adolescents with rheumatic and musculoskeletal diseases (RMD) may be at higher risk for a severe course or worse outcome of COVID-19, and SARS-CoV2 infection may trigger a flare of the RMD. To address these issues, a specific SARS-CoV-2 questionnaire was implemented in the National Pediatric Rheumatology Database (NPRD) in Germany.

**Methods:** Demographic, clinical and treatment data from juvenile patients with RMD as well as data about SARS-CoV-2 infection like test date and method, clinical characteristics, disease course, outcome and impact on the disease activity of the RMD documented on this questionnaire were analyzed.

**Results:** From April 17th, 2020, to February 14th, 2021, data were collected from 79 patients (53% female) with RMD with median age of 14 years, diagnosed with juvenile idiopathic arthritis (57%), autoinflammatory (23%) and connective tissue disease (8%). Sixty-one patients (77%) received disease-modifying antirheumatic drugs (DMARDs), 43% biologic DMARDs, and 9% systemic glucocorticoids. Sixty patients (76%) developed symptoms of COVID-19. Disease severity was mild and outcome was good in the majority of patients. Two patients were hospitalized, one of whom required intensive care and died of cardiorespiratory failure. In 84% of SARS-CoV-2-positive patients, no relevant increase in disease activity of the RMD was observed.

**Conclusions:** In our cohort, COVID-19 in juvenile patients with RMD under various medications was mild with good outcome in the majority of cases. SARS-CoV-2 infection does not appear to have a relevant impact on disease activity of the underlying condition.

## INTRODUCTION

Children and adolescents seem to be less affected by COVID-19 and to have a milder course of COVID-19 than adults ^1^. However, similar to adults, a pre-existing underlying disease or immunosuppression is described as a risk factor for hospitalization in children ^2^. Children and adolescents with rheumatic musculoskeletal disorders (RMD) may be particularly susceptible to SARS-CoV-2 infection, partly because of their underlying disease and also because of therapeutically induced immunosuppression. Furthermore, the immunosuppressive therapy may influence the clinical course of COVID-19 positively or negatively, or that COVID-19 might alter the disease activity of the underlying chronic inflammatory disease.

To address these issues, a specific SARS-CoV-2 questionnaire was implemented into the National Pediatric Rheumatology Database (NPRD) in Germany in 2020 to prospectively gather data i) on clinical characteristics, treatment and outcome of SARS-CoV-2 infection and ii) on the impact of COVID-19 on the pre-existing conditions in juvenile patients with RMD under different immunosuppressive therapies.

Here we report the demographic and clinical characteristics of 79 children and adolescents with RMD and confirmed SARS-CoV-2 infection.

## PATIENTS AND METHODS

The NPRD started 1997 ^3^ and is now being conducted at more than 60 different paediatric rheumatology sites throughout Germany. Yearly, about 15,000 children and adolescents with different RMD, most commonly with juvenile idiopathic arthritis (JIA), but also with connective tissue diseases (e.g., juvenile systemic lupus erythematosus [jSLE], juvenile dermatomyositis [jDM]), vasculitis or autoinflammatory diseases are recorded in the NPRD in a standardized format by disease-specific questionnaires.

The SARS-CoV-2 module of the NPRD was set up on recommendation of the Society for Paediatric Rheumatology (GKJR) in Germany to provide reliable and representative epidemiological data on the frequency, course and outcome of SARS-CoV-2 infection in paediatric patients with RMD. This SARS-CoV-2 module is embedded as a cross-sectional study in the established prospective NPRD and collects information from patients with RMD and all COVID-19 courses (also including asymptomatic SARS-CoV-2 infection), by means of a specific paper-based questionnaire. Next to demographic parameters like age, sex, diagnosis and ethnicity, this questionnaire provides information on the date of a positive SARS-CoV-2 test, method of detection (PCR, antigen test, serology) and reason for testing (symptomatic, contact to a person who tested positive for SARS-CoV-2), on symptoms and clinical manifestations (bronchitis/bronchiolitis, pneumonia, acute respiratory distress syndrome, myocarditis, encephalitis, sepsis, multi-organ failure), the disease course (asymptomatic, mild, moderate, severe, life-threatening defined according to Dong et al. ^4^, laboratory parameters, treatment and outcome of COVID-19.

Furthermore, the drug therapy of the underlying disease at the time of virus detection/diagnosis of COVID-19 (disease-modifying antirheumatic drugs [DMARDs] during the last 6 months, glucocorticoids at time of SARS-CoV-2 infection), changes of treatment (frequency of administration, change in dosage or discontinuation) in response to SARS-CoV-2 infection and the disease activity (21-point numeric rating scale (NRS) 0 – 10, 0 = best) of the underlying disease at the last visit before virus detection and at the corresponding visit of the outcome evaluation are documented.

The introduction of the SARS-CoV-2 module into the NPRD was communicated to all members via email through the GKJR at April 16^th^ 2020, and reporting of appropriate patients was also encouraged through this channel.

This data collection was supplemented by cases of SARS-CoV-2-positive children and adolescents with RMD from the Biologics in Pediatric Rheumatologiy Registry (BiKeR) ^5^, in which SARS-CoV-2 infection was recorded as an “adverse event of special interest”

Additional cases were reported via the COVID-19 Survey of the German Society for Pediatric Infectiology (DGPI, https://dgpi.de/), a registry of hospitalized children and adolescents with SARS-CoV-2 infection.

Only patients with laboratory evidence of SARS-CoV-2 infection were included in our analysis.

The SARS-CoV-2 module of the NPRD was approved by the ethics committee of the Charité - Universitäsmedizin Berlin, as was the data collection of the BiKeR Registry and the DGPI Registry by their respective ethics committees.

### Statistics

Statistical analyses were performed using IBM SPSS Statistics 26 and SAS 9.4M5. Plots were made with Microsoft Excel 2016. Results for continuous variables are presented as medians and interquartile range (IQR) unless otherwise stated. For categorical variables, absolute count is given and proportions of corresponding categories are calculated and presented as percentages. Missing responses were treated on a per-category basis and did not lead to an exclusion of the corresponding case. The difference in the numerical rating scale (Δ NRS), which was used to describe the change of the underlying RMD disease activity, was calculated as the difference of the NRS of the outcome date (NRS-out) and the pre-infection NRS (NRS-pre). Consequently, a non-positive difference describes a constant/decrease of disease activity while a positive difference implies an increase in disease activity. Change in disease activity (“Δ NRS = NRS-out - NRS-pre”) was further categorized as follows: For increase: minimal, if 0 < Δ NRS ≤ 1; minor, if 1 < Δ NRS ≤ 2; relevant if Δ NRS > 2. For decrease: minimal/no decrease: -1 ≤ Δ NRS ≤ 0; minor decrease: -2 ≤ Δ NRS < -1; relevant decrease: Δ NRS < -2. This item was evaluated only in individuals with a date of COVID-19 symptom onset, or – if asymptomatic – with a date of PCR or antigen testing, because these methods allowed to determine the point of time the disease/infection occurred (in contrast to antibody testing)

To determine whether the NRS values were reported in chronological order, i.e. the NRS timeline can be clearly separated into before/after SARS-CoV-2 infection, only valid dates were considered. In cases where only the day was missing, we imputed the missing value with the 15th of the corresponding month.

## RESULTS

### Demographic parameters, diagnostics and treatment

From April 17^th^ 2020 until February 16^th^ 2021, data of 79 patients with RMD were collected. They were predominantly adolescent, and boys and girls were documented with equal frequency. The majority of patients suffered from juvenile idiopathic arthritis (57%), 23% of patients were diagnosed with an autoinflammatory disease (Table 1). In addition to the underlying RMD, 5 patients suffered from asthma, 4 from uveitis, 2 from allergies and 1 patient each from ventricular septal defect, aortic valve insufficiency, Hashimoto’s thyroiditis, hypothyroidism, and diabetes mellitus.

**Table 1:** Demographic parameter, diagnostic tests, diagnoses and treatment of juvenile RMD patients.

| patients | Total cohort | Symptomatic | Asymptomatic |
| --- | --- | --- | --- |
| Total | 79 | 60 | 19 |
| Female, % | 53 | 54 | 46 |
| Age, median (IQR) | 14 (11 – 16) | 14 (12 – 17) | 12 (7 – 16) |
| **Test method, n (%)** |  |  |  |
| PCR | 52 (66) | 39 (65) | 13 (68) |
| Antigen test | 3 (4) | 3 (5) | 0 (0) |
| Antibody test | 19 (24) | 14 (23) | 5 (26) |
| Laboratory test not specified | 5 (6) | 4 (7) | 1 (5) |
| **Juvenile idiopathic arthritis, n (%)** | 45 (57) | 33 (55) | 12 (63) |
| Oligoarthritis (persistent/extended) | 16 | 11 | 5 |
| Polyarthritis (RF negative/ positive) | 18 | 15 | 3 |
| Enthesitis-related arthritis | 7 | 4 | 3 |
| Systemic juvenile idiopathic arthritis | 3 | 3 | 0 |
| Other arthritis | 1 | 0 | 1 |
| **Autoinflammatory diseases, n (%)** | 18 (23) | 18 (30) | 0 |
| FMF | 11 | 11 | 0 |
| NLRP3-associated autoinflammatory disease | 2 | 2 | 0 |
| PFAPA | 1 | 1 | 0 |
| TRAPS | 3 | 3 | 0 |
| DADA2 | 1 | 1 | 0 |
| **Connective tissue disease, n (%)** | 6 (8) | 3 (5) | 3 (16) |
| SLE | 3 | 2 | 1 |
| JDM | 2 | 1 | 1 |
| Scleroderma (localized) | 1 | 0 | 1 |
| **Other, n (%)** | 10 (13) | 6 (12) | 4 (21) |
| M. Behcet | 1 | 1 | 0 |
| CRMO | 5 | 3 | 2 |
| SpA* | 1 | 1 | 0 |
| Sjogren's syndrome | 1 | 1 | 0 |
| Uveitis (idiopathic) | 2 | 0 | 2 |
| **Treatment, n (%)** | **69 (87)** | **51 (85)** | **18 (95)** |
| Systemic glucocorticoids | 8 | 4 | 4 |
| any DMARD | 60 | 49 | 11 |
| MTX | 19 | 15 | 4 |
| bDMARD | 33 | 28 | 5 |
| TNF-alpha antagonist | 20 | 17 | 3 |
| MTX+TNF-alpha antagonist | 4 | 4 | 0 |
| IL-6 blockade | 6 | 5 | 1 |
| IL-1 blockade | 4 | 4 | 0 |
| Baricitinib | 1 | 1 | 0 |
| ...Colchicine | 9 | 8 | 1 |
| ...Hydroxychloroquine | 7 | 4 | 3 |
| Mycophenolate mofetil | 3 | 2 | 1 |
| Sulfasalazine | 1 | 1 | 0 |
| ...Azathioprine | 2 | 2 | 0 |

**Abbreviations:** PCR: polymerase chain reaction; RF: rheumatoid factor; FMF: Familial Mediterranean Fever, PFAPA: periodic fever, aphthous stomatitis, pharyngitis, cervical adenitis; TRAPS: TNF-receptor-associated periodic syndrome; DADA2: Deficiency of adenosine deaminase 2; SLE: Systemic lupus erythematosus, JDM: Juvenile dermatomyositis; CRMO: Chronic recurrent multifocal osteomyelitis; SpA: Spondylarthritis, the patient was older than 16 years at the time of diagnosis and therefore could not be classified as having enthesitis-related arthritis; (b)DMARD: (biological) disease modifying antirheumatic drug; MTX: methotrexate

Most patients were tested for SARS-CoV-2 because they had contact with an infected person (52/79, 66%); only 26 (33%) were tested because of typical symptoms. The preferred test method was PCR (66%), followed by serology testing (24%) and antigen detection (4%); in 5 patients the laboratory method was unknown.

Sixty patients (76%) were treated with a DMARD at the time of SARS-CoV-2 infection, and 42% with a biologic DMARD.

DMARD therapy (9x bDMARD, 2x bDMARD+MTX, 3x MTX only) was discontinued or changed in frequency of application and/or dosage in response to confirmed SARS-CoV-2 infection in 14 patients with the following diagnoses and treatments: seronegative polyarthritis (PA) n=7, enthesitis-related arthritis n=2, seropositive PA, oligoarthritis (OA) persistent, OA extended, chronic recurrent multifocal osteomyelitis (CRMO), deficiency of adenosine deaminase 2 each n=1.

### Clinical characteristics of SARS-CoV-2 infection in juvenile patients with RMD

Sixty patients (76%) developed symptoms of COVID-19, 19 patients (24%) remained asymptomatic.

All patients with autoinflammatory diseases as well as all patients with systemic JIA - even though only 3 patients - developed symptoms of COVID-19 whereas 29% of patients with the other forms of JIA and 50% of patients with connective tissue diseases remained asymptomatic.

The clinical symptoms of COVID-19 in the 60 symptomatic juvenile patients with RMD are summarized in figure 1. The most common symptom of SARS-CoV-2 infection was fever (40%), although usually not high: of the 14 patients who provided information on fever level, only one had a body temperature > 39.5°C; 50% reported a temperature in the range of 38.5°C and 39.5°C and 32 % reported an elevated temperature < 38.5°C.

**Figure 1:**
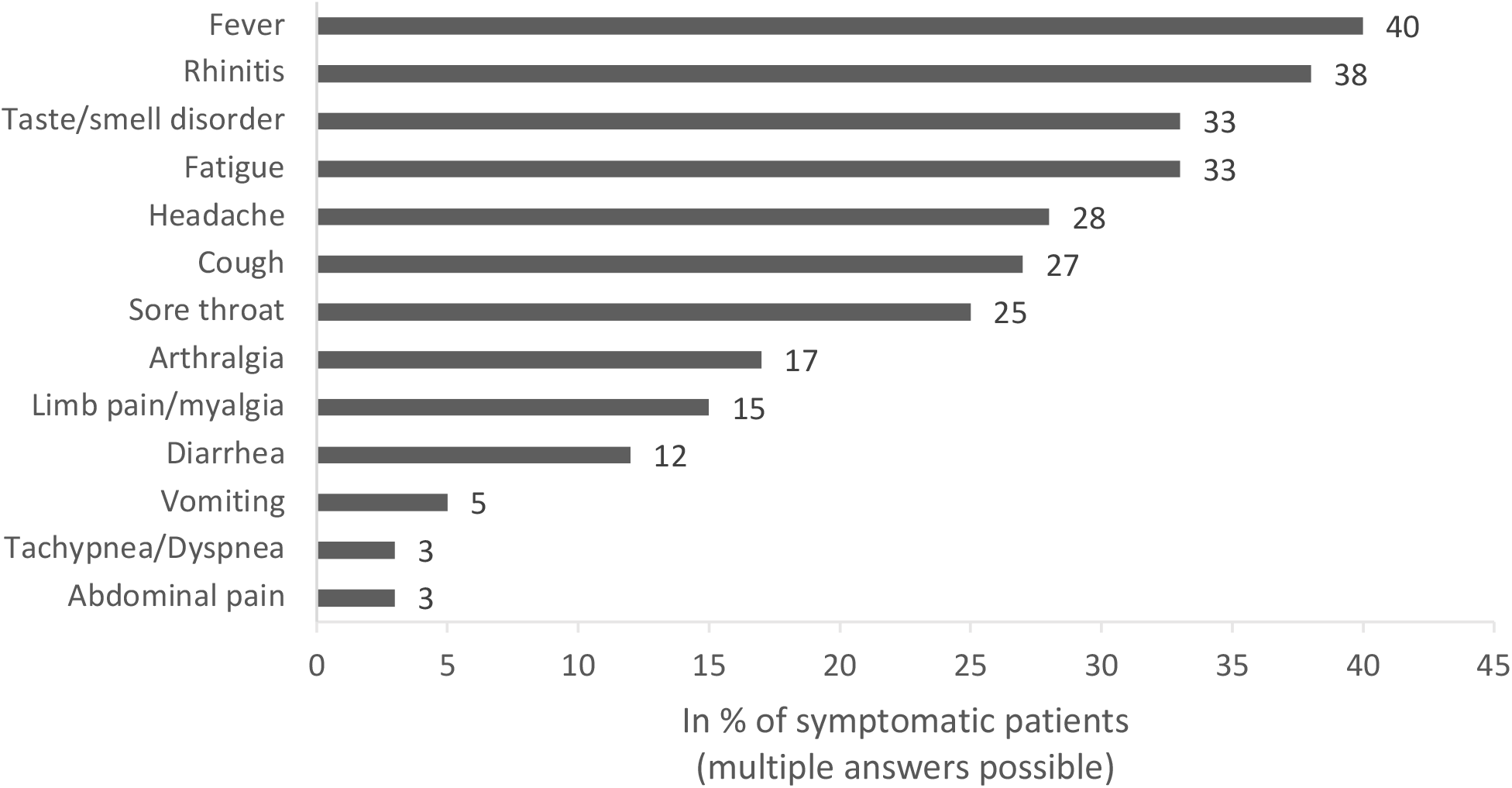
Clinical symptoms of SARS-CoV-2 infection in 60 juvenile patients with RMD

One patient who was hospitalized was diagnosed with pneumonia, but no outpatients (n=77) were reported to have pneumonia; 2 outpatients were diagnosed with bronchitis/bronchiolitis.

Due to the small number of documented laboratory results, we were unable to perform any analyses in this regard.

### Course and Outcome of COVID-19 in juvenile patients with RMD

Disease course was classified as mild in 46 of 48 symptomatic patients (96%) with valid data on this item. None of the symptomatic patients with missing data regarding the disease course of COVID-19 (n=12) were hospitalized.

Two patients were reported via the DGPI registry for hospitalized children with COVID-19: A girl with chronic nonbacterial osteitis on therapy with NSAID was hospitalized but did not show clinical signs of pneumonia nor required oxygen administration or intensified drug therapy. The second patient was a girl who was initially classified as having systemic JIA at the age of 1 year and then as polyarticular JIA in the further disease course, and treated with methotrexate as well as prednisolone. After having experienced a second febrile seizure she was diagnosed with sinus vein thrombosis and cerebral edema on computer tomogram. This patient died despite intensive care 8 days after symptom onset. The cause of death was considered cardiorespiratory failure with radiographic changes in chest X-ray compatible with COVID-19. A genetic examination revealed a so far unrecognized immunodeficiency.

Of the 52 symptomatic patients for whom COVID-19 outcome information was available, 41 patients (79%) reported that their health status was recovered (median time from symptom onset to outcome evaluation 28 days (Interquartile range (IQR) 9 – 55), 10 patients (19%) stated that their health status had not yet recovered after COVID-19, mostly due to a continuing taste/smell disorder (mean time between symptom onset and outcome evaluation was 26 days, IQR 7 – 47).

### Outcome of the underlying RMD after SARS-CoV-2 infection

We were able to evaluate the disease activity of the underlying RMD (NRS) before and after the proven SARS-CoV-2 infection in 32 individuals (28 symptomatic and 4 asymptomatic patients). The remaining patients in our study lacked the necessary information to do so. The median time from the last assessment of RMD disease activity of the chronic inflammatory disease to positive SARS-CoV-2 testing was 52 days (IQR 37.5-102.5 days). After detection of SARS-CoV-2 infection, the median time to outcome assessment of RMD disease activity was 31 days (IQR 13 – 50 days). The mean disease activity with a range of 0 (= best) to 10 of those 32 patients was 0.90 ± 1.37 (median 0.5, IQR 1 – 1.25) before SARS-CoV-2 infection and 1.14 ± 2.12 (median 0.25, IQR 0 – 1.25) at the time of outcome assessment. No up to minimal increase in disease activity (difference in NRS before versus after SARS-CoV-2 infection ≤ 1) of the underlying disease was recorded in 27 of 32 patients (84%), regardless of whether patients were asymptomatically infected or developed COVID-19 (Figure 2). An increase in disease activity with a difference in NRS before versus after SARS-CoV-2 infection > 1 was seen in 5 patients (4 symptomatic, 1 asymptomatic).

**Figure 2:**
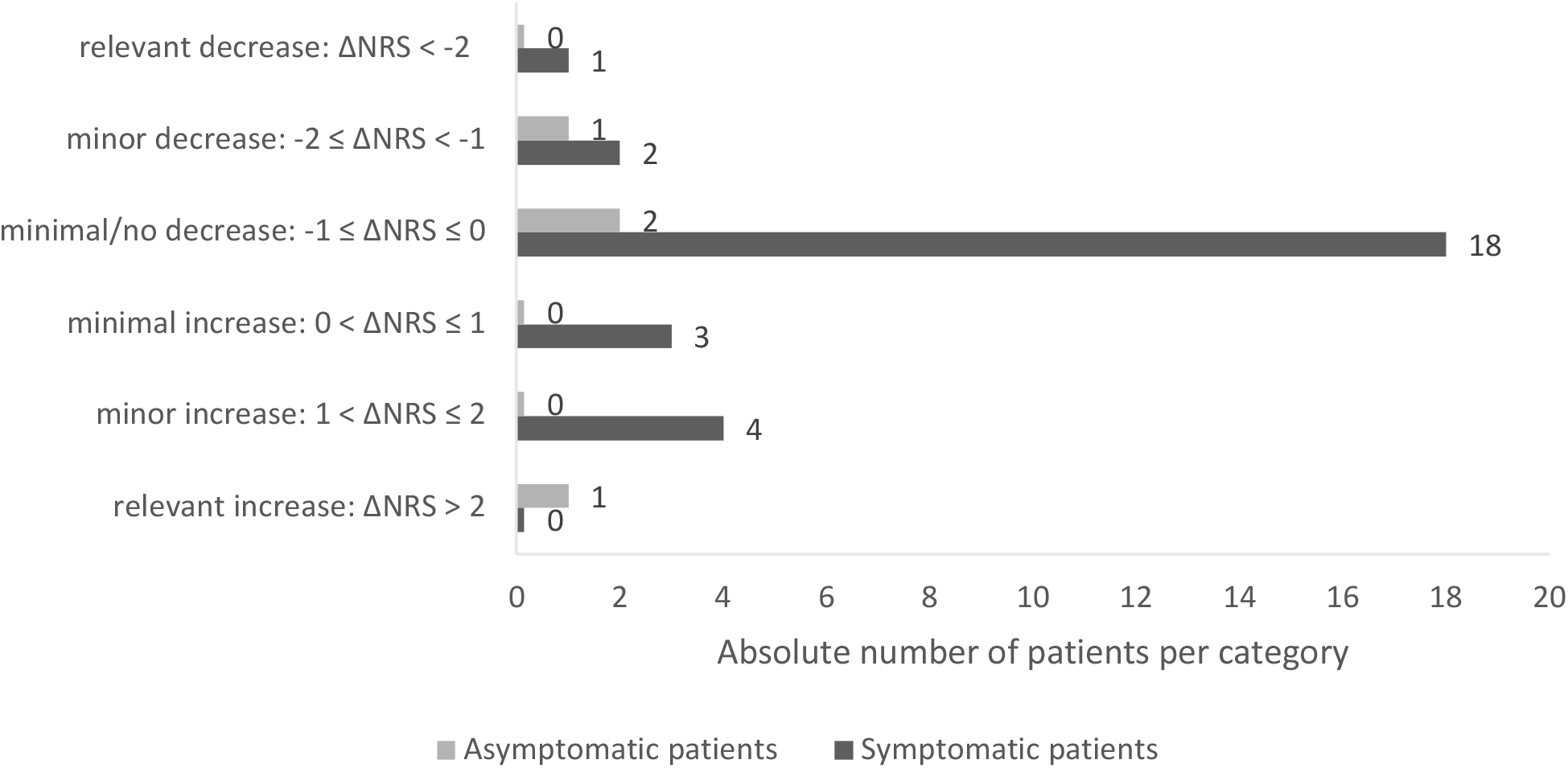
Changes in NRS disease activity of the underlying rheumatic disease before versus after SARS-CoV-2 infection

All but one (n=13) of the patients in whom drug therapy had been discontinued or the dosing/interval of application of medication had been changed had an NRS of less than or equal to 1 with respect to disease activity of the underlying disease before COVID-19, which changed by a maximum of 0.5 thereafter. One patient, who had paused tocilizumab for 2 doses, experienced a flare of his seronegative polyarthritis (NRS-out: 9) 2 months after asymptomatic SARS-CoV-2 infection (NRS before infection was 3).

Another patient experienced a recurrence of uveitis 8 weeks after symptom onset of COVID-19 without a flare of her oligoarthritis while maintaining DMARD therapy with MTX.

## DISCUSSION

Although children and adolescents are known to develop COVID-19 less frequently and usually more mildly than adults ^1 4 6^, the clinical characteristics of COVID-19 in terms of symptoms, severity of course and outcome, as well as the impact of this infection on the underlying disease in juvenile patients with rheumatic or autoinflammatory diseases, has remained unclear.

In the study presented here, the majority of patients diagnosed with RMD under different therapeutic regimens with confirmed SARS-CoV-2 infection have shown a mild disease course with good outcome. Only two patients were hospitalized, of whom one patient had a severe course of COVID-19 with a fatal outcome.

Of the total 79 patients in our cohort, 24% were asymptomatically infected and 76% showed symptoms of illness from COVID-19. Fever and rhinitis were most commonly reported (40% and 38% respectively), followed by fatigue, loss of smell and/or taste (33%), headache, sore throat and cough. This frequency distribution of symptoms found here is largely consistent with published data on the clinical manifestation of COVID-19 in children and adolescents in the general population^167^. However, we have a relatively high proportion of SARS-CoV-2 positive patients in our cohort who reported smell and/or taste disturbances, which has been identified as a fairly specific sign of SARS-CoV-2 infection ^8^.

In our cohort, the disease course of COVID-19 was mild in 96% of patients with valid data for this item; none of the patients who lacked formal information on disease course was hospitalized, so it may be concluded that these patients at least did not have a severe course of COVID-19. Due to this rather homogeneous result in the evaluation of COVID-19 disease course and the low number of hospitalized patients, the influence of comorbidities or drug therapy of the underlying disease on the expression of SARS-CoV-2 infection could not be analyzed. From the National Registry of Adult Patients with Rheumatic Diseases and SARS-CoV-2 Infection ^9^ in Germany, it was reported that glucocorticoid therapy but not DMARD therapy was associated with a higher risk of hospitalization related to COVID-19. DMARDs may even prevent a severe COVID-19 course, as they also attenuate immune processes triggered by the viral disease, including an exaggerated immune response such as the so-called ‘cytokine storm.’ Studies on the efficacy of antirheumatic therapeutics in severe COVID-19 courses showed partly conflicting ^10-13^, partly promising results ^14^.

Filocamo et al ^15^ used a survey to collect data from 123 children and adolescents with chronic rheumatic disease treated with a bDMARD in Milan, Italy, during the first wave of SARS-CoV-2 infection. Eight children presented with mild respiratory symptoms, three of whom had an adult in the family with suspected SARS-CoV-2 infection; however, no such infection was detected in any of the children.

Koker et al ^16^ surveyed parents of 414 children and adolescents with rheumatic diseases and immunosuppressive treatment by telephone interview regarding possible COVID-19-associated symptoms, risk contacts, performance of diagnostic tests (SARS-CoV-2-PCR), and possible interruption of antirheumatic therapy, and recorded clinical parameters and therapies from medical records (7). At the time of the survey, 42.3% were receiving bDMARDs, 28.7% nbDMARDs, and 14.5% both; a quarter of the cohort had comorbidities. Nine patients were screened in hospital for COVID-19, of whom 6 had close contact with a confirmed infectious case; only one patient was found to have SARS-CoV-2 infection (with an uncomplicated course).

Thus, juvenile patients with rheumatic diseases on DMARD therapies do not appear to be more likely to be infected with SARS-CoV-2 than children and adolescents in the general population. Possibly, they protect themselves through appropriate behavior (distance requirement, hygiene rules), as it was shown for adult patients with RMD ^17^. Similarly, in studies by Michelena et al ^18^ from Spain and by Favalli ^19^ from Italy, the incidence of COVID-19 in adult patients with rheumatic diseases on therapy with biologics and synthetic DMARDs was comparable to the rate in the general population.

Some studies investigated the incidence of hospitalization - as a surrogate parameter for a more severe course of COVID-19 - in adult patients with rheumatic diseases compared with the normal population: Haberman et al. ^20^ reported a similar rate of hospitalization of patients with RMD in the setting of SARS-CoV-2 infection compared with the normal population in New York City ^20^.

Furthermore, based on the data in this study, they concluded that therapy with bDMARD or tsDMARD was not associated with a worse outcome for COVID-19 in patients with RMD. Jovani et al. reported an estimated odds ratio of 2.61 for hospitalization of patients with RMD on bDMARD/tsDMARD but found no elevated risk of developing severe COVID-19 in these patients compared with healthy subjects ^21^. Nevertheless, some studies actually showed evidence of a more severe course of COVID-19 in patients with rheumatic diseases: D’ Silva et al. ^22^ compared manifestation and outcome of COVID-19 in patients with and without RMD and found similar symptoms and laboratory changes as well as comparable hospitalization and mortality rates in both patient groups. However, patients with RMD were more likely to need intensive care and also mechanical ventilation (multivariable OR 3.11 with 95% confidence interval 1.07 to 9.05 ^22^). A higher rate of respiratory failure in the context of COVID-19 compared with the general population was also found by Ye et al. in their study of 21 adult patients with RMD in Wuhan, China ^23^.

Recently, using registry data from Denmark, Cordtz et al. ^24^ reported that adult patients with inflammatory rheumatic diseases were both more frequently hospitalized and at higher risk of severe COVID-19 (as measured by the number of patients with intensive therapy, ARDS, or death ^24^. Differences in the composition of the patient populations with respect to age, underlying disease (rheumatoid arthritis, ankylosing spondylitis, connective tissue disease, vasculitides, systemic autoimmune disease, etc.), disease duration and comorbidities as well as therapies might explain these discrepant findings.

In our study with juvenile patients with RMD, 96% had a mild disease and the overall outcome of COVID-19 in these patients with defined chronic rheumatic or autoinflammatory diseases was also good with health condition restored in 79% of patients four weeks after symptom onset of COVID-19. None of the outpatients was diagnosed with pneumonia, only 2 patients were hospitalized. One patient died in whom next generation sequencing revealed a so far unrecognized underlying immunodeficiency that was presumably responsible for the fatal course.

In 84% of our patients, there was no or only minimal increase in disease activity of the underlying RMD (increase in NRS ≤ 1). One patient with comorbid anterior uveitis suffered a uveitis recurrence after COVID-19 without a flare of her oligoarthritis while maintaining DMARD therapy with MTX. In contrast, one patient with seronegative polyarthritis experienced a flare of his PA after discontinuation of tocilizumab in the setting of asymptomatic SARS-CoV-2 infection. This patient, unlike all other patients who had DMARD therapy reduced or discontinued after diagnosis of COVID-19 with no increase of disease activity thereafter, had an NRS of 3 (moderate disease activity) already prior to his SARS-CoV-2 infection. Here, the pause in therapy rather than the SARS-CoV-2 infection (especially as asymptomatic) seems to be causative for the flare, particularly since the patient was not in remission before this treatment stop.

### Limitations

Our data collection was based on voluntary reporting of confirmed SARS-CoV-2 infections in juvenile patients with RMD and therefore likely does not include all such patients in Germany. The relatively small number of cases of each disease entity and the variety of therapies do not allow reliable conclusions regarding implications of a specific rheumatic disease or the effects of medication on the clinical expression and outcome of SARS-CoV-2 infection in these patients.

Assessment of disease activity before and after SARS-CoV-2 infection was performed as part of routine clinical practice and not after a fixed interval. Therefore, it is possible that flares that occurred in the period after SARS-CoV-2 infection but before outcome assessment were not recorded. Furthermore, we did not have a comparison group of SARS-CoV-2-negative patients with RMD regarding the change in NRS of disease activity over a comparable period of time.

### Summary

In summary, the clinical characteristics of COVID-19 in this patient group are similar to those of healthy peers; disease course was mild with and without maintaining DMARD therapy and COVID- 19 outcome was good in most of these patients. Furthermore, SARS-CoV-2 infection does not appear to have a major effect on the underlying disease activity, whereas discontinuation of therapy might pose a risk of flare in patients with moderately active disease.

To our knowledge, our cohort currently represents the largest collection of data from children and adolescents with RMD and confirmed SARS-CoV-2 infection on various immunomodulatory therapies. Further studies with larger numbers of cases are needed to better investigate associations between the underlying disease, its drug therapy, and the clinical expression and possible consequences of a SARS-CoV-2 infection in these patients.

## Data Availability

All data relevant to the study are included in the article.

## Acknowledgements

We are grateful to all patients and their parents for participating in the NRPD, BiKeR and the COVID-19 Survey of the DGPI and to all colleagues who contributed to this data collection.

Dr. Annett Lambrecht, Universitätskinderklinik Magdeburg

Dr. Caroline Siemer, DZKJR Garmisch-Partenkirchen

Dr. Ralf Trauzeddel, Helios-Klinikum Berlin-Buch

Dr. Moritz Klaas, Vivantes Klinikum Berlin-Friedrichshain PD

Dr. Jürgen Brunner, Universitätsklinderklinik Innsbruck

Dr. Gonza Ngoumou, Universitätsmedizin Berlin - Charité

Dr. Mareike Lieber, Universitätsmedizin Berlin - Charité

Dr. Sae Lim von Stuckrad, Universitätsmedizin Berlin - Charité

Dr. Lisa Lurz, Universitätsmedizin Berlin - Charité

Dr. Peggy Rühmer, Helios Kliniken Vogtland

Dr. Andrea Skrabl-Baumgartner, Universitätskinderklinik Graz Prof.

Dr. Tim Niehues, Helios Kliniken Krefeld

PD Dr. Catharina Schütz, Universitätskinderklinik Dresden

Dr. Anja Hauenherm, Städt. Kliniken St. Georg, Leipzig

Dr. Friederike Blankenburg, Klinikum Stuttgart

Dr. Anita Heinkele, Klinikum Stuttgart

Dr. Frank Dressler, Medizinische Hochschule Hannover

Dr. Tanja Hinze, Universitätskinderklinik Münster

Dr. Sebastian Schua, St. Josef Stift Sendenhorst

Prof. Dr. Markus Hufnagel, Universitätskinderklinik Freiburg

Dr. Melanie Römer, Kinderarztpraxis, Kempten

Dr. Veit Grote, Dr. von Hauner‘sches Kinderspital, LMU München

Dr. Regine Borchers, Universitätsklinikum Augsburg

## Contributors

KM, CS, TK developed the study concept and design. MN was responsible for the administrative and technical support The funding was obtained by KM, GH and JA. Data were acquired by KM, GH, JBD, CS, RB, HG, RH, MB, AH, WE, JA, AK and TK. Data were analyzed and interpreted by SE, CS and KM. CS and SE drafted the manuscript. All critically revised the manuscript and approved the final submitted draft.

## Ethics approval

The NPRD was approved by the ethics committee of the Charité - Universitätsmedizin Berlin (EA1/044/07). The Biologics in Pediatric Rheumatology Registry (BiKeR) was approved by the ethics committee of the physician board Aerztekammer Nordrhein, Duesseldorf (reference number 2/2015/7441). The COVID-19 Survey was approved by the Ethics Committee of the Technische Universität (TU) Dresden (BO-EK-110032020)

Parents and patients from the age of 8 years on gave their informed assent/consent for participation. The procedures used in this study adhere to the tenets of the Declaration of Helsinki.

